# Common genetic determinants of dynamic human in vivo immune response to *Mycobacterium tuberculosis*

**DOI:** 10.64898/2026.07.06.26357408

**Authors:** Ping Zhang, Carolin T Turner, Aneesh Chandran, Tom Parks, Joshua Rosenheim, Jana Jiang, Lucy CK Bell, Rubin Rose-Key, Michelle Berkeley, Santino Capocci, Marc Lipman, Heinke Kunst, Stefan Lozewicz, Gillian S Tomlinson, Julian C Knight, Mahdad Noursadeghi

## Abstract

We investigated host-genetic TB susceptibility by expression quantitative trait loci (eQTL) analysis of the tuberculin skin test (TST) as a standardised challenge model of human *in vivo* TB immunology. Paired genotyping with 415 RNA-sequencing profiles from day 2 and day 7 TST biopsies in 267 individuals with latent or active TB identified cis-eQTLs affecting 1,719 response genes. The strongest signal mapped to *ERAP2*, and colocalisation analysis linked reduced *ERAP2* expression to increased TB risk in GWAS data. Heritability was greatest in HLA class II antigen presentation and T-cell activation pathways. HLA-DR haplotypes associated with subsequent expansion of Mtb-reactive T cells, linking host genotype to antigen-specific immunity. Trans-eQTLs also identified a proliferative programme centred on *NCAPD3*, implicating genetically regulated cell-cycle control as an antigen-independent determinant of T-cell immunity. These findings provide a functional framework for interpreting TB susceptibility loci and identification of candidate biomarkers for TB risk stratification and vaccine development.

## Introduction

Most incident infections do not lead to tuberculosis (TB) disease, but the mechanisms that determine different clinical outcomes of infection are not known. Immune responses are thought to be important. CD4 T cells, macrophages, and the cytokines interferon (IFN)γ and tumour necrosis factor (TNF) in particular are thought to be necessary for protective immunity^1–5^, but deficiencies in any of these responses do not explain incident disease in the vast majority of cases within the general population.

There is evidence of population level genetic susceptibility to TB disease in twin studies, in which monozygotic twin contacts of index TB cases have approximately 3-fold higher risk of TB disease compared to dizygotic twins^6^. However, genome-wide association studies (GWAS) have failed to show statistically robust and reproducible associations with disease^7^. Common genetic variation represented by single nucleotide polymorphisms (SNPs) in regulatory sequences are widely recognized as a prevalent source of quantitative variation in gene expression including variation in transcriptional upregulation of immune response genes^8^. Identification of immune response gene expression quantitative trait loci (eQTLs) has been almost exclusively limited to ex vivo stimulation of selected immune cell populations, and yet to be validated within complex dynamic multicellular *in vivo* immune responses, notably within a disease relevant context.

We established transcriptional profiling of the tuberculin skin test (TST) as a standardised human experimental challenge to investigate variation of *in vivo* immune responses to *Mycobacterium tuberculosis* (Mtb) between groups of individuals. We have previously shown that transcriptional perturbation in the immune response to the TST captures all the variance in transcriptional profiles of tissue from the site of lung TB granuloma, providing strong evidence that the TST comprises the full repertoire of molecular changes at the natural site of disease^9^. We have used this model to evaluate differences associated with disease^10,11^, HIV co-infection^9^ and the impact of anti-TNF therapy^12^. Single cell sequencing analysis of this response indicated the TST is dominated by recruitment of T cells, NK cells and monocyte derived cells^13^. T cell receptor sequencing analysis revealed an influx of polyclonal non-antigen specific T cells at the time of maximal inflammatory induration after 2 days that is replaced by oligoclonal expansion of Mtb reactive T cells at 7 days^14^.

In the present study, we sought to evaluate inter-individual variation at the level of gene expression in day 2 and day 7 TSTs and identify the SNP eQTL that may underpin variation in expression of local likely cis-regulated genes. We investigated the dominant immune pathways that were subject to cis-eQTL regulation. We used *in silico* approaches to identify the most likely causal SNPs, the cellular context in which they exert their effects, the gene regulatory mechanisms that mediate control of gene expression, and used colocalization analysis to investigate the association of TST eQTL with disease traits in GWAS data. Finally, in view of the critical role of T cell responses in TB immunology, we explored the genetic determinants of the temporal evolution of Mtb-reactive T cell responses.

## Results

### Interindividual variation in TST immune responses at the molecular level

The TST has long been used as a quantitative *in vivo* measure of T cell reactivity to Mtb antigens with a high level of concordance to peripheral blood interferon (IFN)γ production by antigen stimulated T cells^15^. We have shown that transcriptional profiling of skin biopsies from the TST site reveals enrichment of wide-ranging transcripts reflecting the functional activity of canonical innate and adaptive immune pathways in response to Mtb challenge. We have previously focussed on variation in the TST transcriptome at the pathway level, to identify between-group differences in human *in vivo* immune responses associated with protective or pathogenic immunity^9–11^, and temporal evolution of maximal inflammatory responses in the day (D)2 TST to local enrichment of Mtb reactive T cell repertoire in the D7 TST^14^.

In the present study we evaluated within-group inter-individual variation at the level of individual gene expression in data sets obtained from D2 and D7 TSTs in a cohort of healthy people with Mtb infection labelled latent TB infection (LTBI), and D2 TSTs from a cohort of patients with active TB (ATB) disease within 28 days of diagnosis and treatment initiation (Supplementary Figure 1). In each group, we defined the TST response transcriptome by significantly higher transcript abundance compared to transcriptomes from similar skin biopsy samples at the site of control saline injections, reported previously^10^. Variation in gene expression of the TST transcriptome ranged from a standard deviation of 0.2-6.8 in Log2 transcript abundance, with an average standard deviation of 1.2 Log2 transcript abundance for D2 TSTs and D7 TSTs in LTBI, and standard deviation of 1.0 Log2 transcript abundance for D2 TSTs in ATB. Consistent with the prevailing evidence that the TST is dominated by IFNγ-dependent responses, the first principal component of the TST transcriptome was strongly correlated with IFNγ gene expression. However, subsequent principal components of the transcriptional data showed diminishing correlation with IFNγ (Supplementary Figure 2). Moreover, at the level of individual genes, we found substantial variation in interindividual gene expression and correlation with *IFNG* expression, revealing IFNγ-independent regulation of immune responses (Figure 1A).

**Figure 1.**
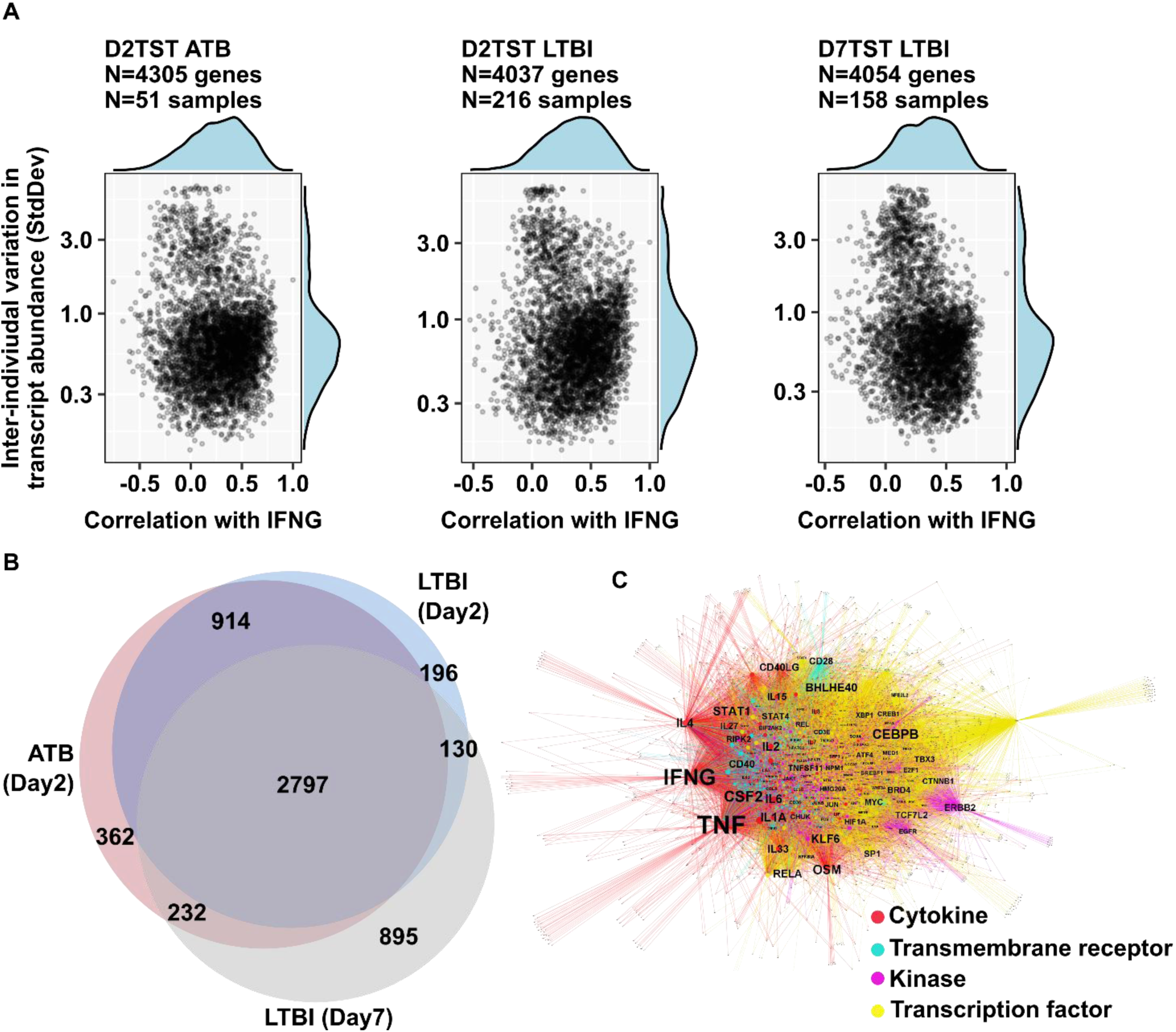
Inter-individual variation of TST response genes involved in canonical pathways of cell-mediated immunity. **(A)** Inter-individual variation is shown as standard deviation of log2-transformed expression levels (tpm) for significantly enriched gene transcripts in TST response transcriptomes and their correlation with *IFNG* gene expression. Data are from three bulk RNA sequencing data sets: day 2 TSTs from individuals with active tuberculosis (ATB) disease, day 2 TSTs from individuals with latent TB infection (LTBI), and day 7 TSTs from individuals with LTBI. **(B)** Venn diagram comparing TST response genes between the three data sets. **(C)** Network plot of predicted upstream regulators (labelled nodes) and their putative gene targets in the TST response transcriptome (unlabelled nodes), stratified in colour by functional annotations based on analysis of the integrated TST response genes from each of the three data sets.

### Identification of genome-wide expression quantitative trait loci (eQTL) for the TST transcriptome

Next, we used the TST model to test the hypothesis that the gene-level inter-individual variation of *in vivo* immune responses to Mtb is associated with common genetic variation, due to SNPs in putative regulatory regions that may impact quantitative gene expression. We sought to maximise statistical power to identify eQTLs by focussing on local likely cis associations within 1MB of the transcriptional start site for each gene in TST transcriptomes across the combined LTBI and ATB cohorts of 134 male and 140 female individuals with diverse ethnicities (Table 1). We found extensive overlap between the expressed genes for TST transcriptomes of the D2 TST in ATB, D2 TST in LTBI and D7 TST in LTBI datasets (Figure 1B). Therefore, in our primary analysis, we used the combined dataset representing a diverse range of immune pathways (Figure 1C, Supplementary Table S1) from 415 TST samples derived from 267 individuals for which we had paired genotyping data in order to maximise statistical power for identification of cis-acting QTL.

**Table 1.**

|  | ATB (2 day TST) | LTBI (All participants) |
| --- | --- | --- |
| **Total** | 51 | 223 |
| **Sex** |  |  |
| Female | 20 (39%) | 120 (54%) |
| Male | 31 (61%) | 103 (46%) |
| **Ethnicity** |  |  |
| African | 12 (24%) | 66 (30%) |
| American | 1 (2.0%) | 3 (1.3%) |
| East Asian | 5 (9.8%) | 41 (18%) |
| European | 21 (41%) | 65 (29%) |
| Mixed | 2 (3.9%) | 1 (0.4%) |
| South Asian | 10 (20%) | 43 (19%) |
| NA | 0 (0%) | 4 (1.8%) |
| **Age** | 30 (27, 43) | 34 (29, 43) |

| LTBI (2 day TST) | LTBI (7 day TST) |
| --- | --- |
| 216 | 158 |
| 115 (53%) | 85 (54%) |
| 101 (47%) | 73 (46%) |
| 64 (30%) | 48 (30%) |
| 3 (1.4%) | 2 (1.3%) |
| 40 (19%) | 27 (17%) |
| 64 (30%) | 44 (28%) |
| 1 (0.5%) | 1 (0.6%) |
| 40 (19%) | 34 (22%) |
| 4 (1.9%) | 2 (1.3%) |
| 34 (29, 43) | 35 (29, 45) |

<sup>1</sup>n (%); Median (Q1, Q3)
ATB = individuals with active TB disease
LTBI = individuals with latent TB infection
TST = tuberculin skin test

We identified 7411 genes showing likely cis-acting SNP associations with gene expression (referred to as eGenes) across the genome after correcting for linkage disequilibrium (LD) between SNPs using eigenMT^16^ (Supplementary Table S2). Of these, 23.2% (1719 of eGenes) were also TST response genes (Figure 2A). The strongest cis-eQTL association was evident for the Endoplasmic Reticulum Aminopeptidase (*ERAP*2) gene, the protein product of which is best known for peptide processing in the HLA class 1 antigen presentation pathway^17^. Through conditional analysis, we identified a total of 9119 independent associations (Supplementary Table S3), with multiple signals detected for 1439 TST eGenes, including a median of 2 and a maximum of 7 conditional cis-acting SNP association signals per gene. For each eGene, the lead SNP was defined as the cis-variant showing the strongest association (lowest nominal p-value) with gene expression.

**Figure 2.**
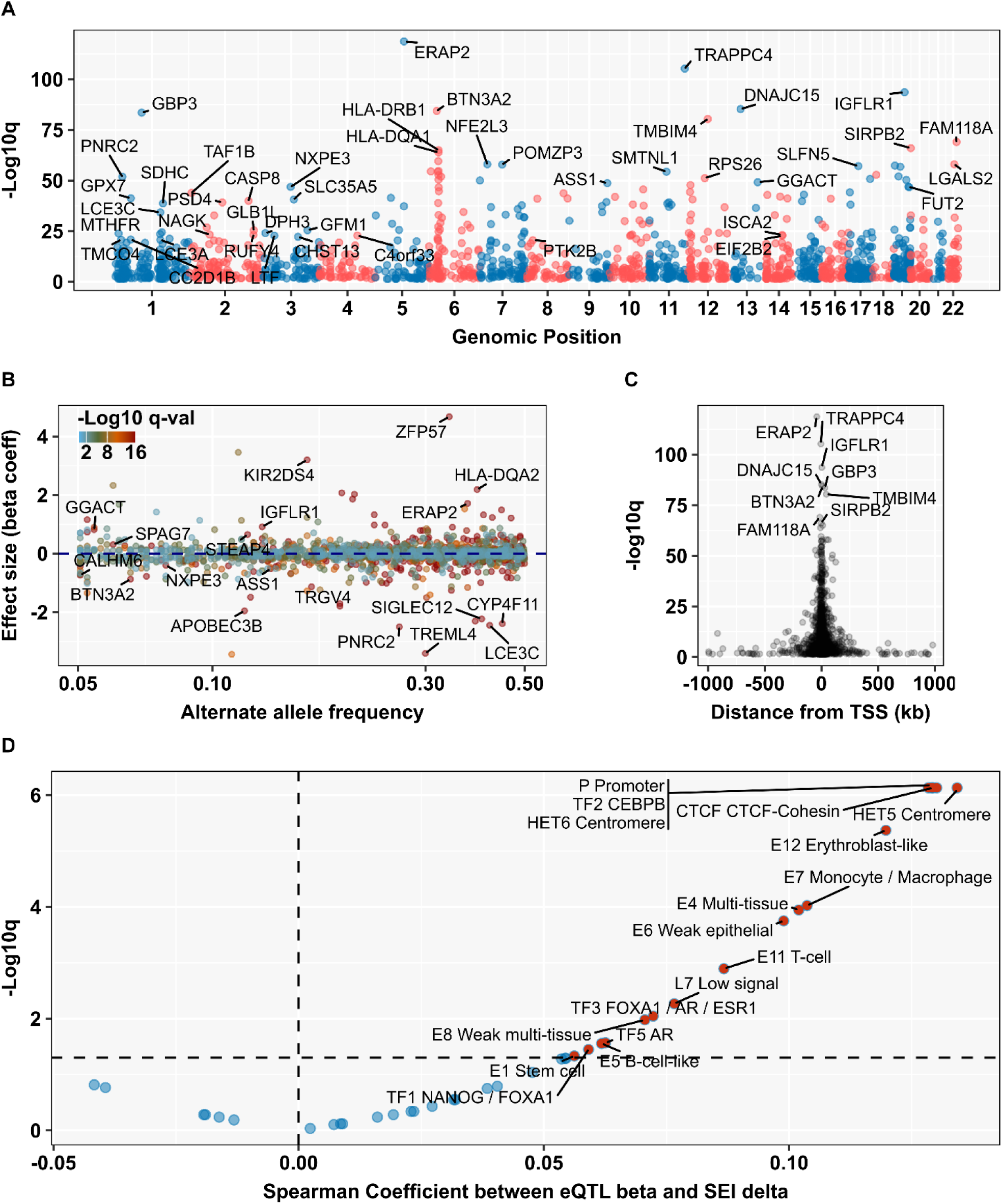
Genome-wide TST response cis-QTL and relationship to gene regulatory sequences. **(A)** Manhattan plot of lead cis-QTL for TST response genes in integrated TST transcriptomes showing chromosomal location (alternating red/blue label) and statistical confidence in their associations with the cis-gene expression represented as -Log10 transformed false discovery rates (q value). **(B)** Effect size (represented by the beta coefficient) and alternate allele frequency and **(C)** Distance from transcriptional start site of lead cis-QTL for TST response genes, stratified by statistical confidence in their associations with cis-genes. Labels in A-C indicate selected cis-genes among TST response cis-QTL. **(D)** Strength of statistical correlations between the effect size of lead cis-QTL for TST response genes and probability of their association with regulatory sequence clusters derived functional genomics data as assessed using the Sei framework (https://humanbase.flatironinstitute.org/sei)^18^. Labels in panel D indicate selected regulatory ‘sequence classes’ derived from the Sei database. Red nodes represent statistically enriched regulatory sequence classes.

Across the range of alternative allele frequencies for the lead cis-eQTL, the beta coefficient for the effect of alternative alleles on log2 eGene expression ranged from -3.4 for *SIGLEC14* to 4.7 for *ZFP57* (Figure 2B, Supplementary Figure 5A). A relationship between eQTL frequency with alternate allele frequency was evident, most likely reflecting the limitations of statistical power in our sample size (Supplementary Figure 5B). In addition, there was a strong relationship between statistical confidence in the lead cis-eQTL SNPs and proximity to the transcriptional start site for the eGene consistent with the hypothesis that these cis-eQTL are predominantly situated in gene promoter sequences (Figure 2C).

### Functional mechanisms and cellular context of TST cis-QTL

To build confidence in the causal role of our lead cis-eQTL SNPs, we explored their putative functional effects *in silico* using deep learning models derived from collated functional genomics data^18^. These models assign probabilities for the association of given sequences with regulatory sequence clusters for specific regulatory functions. Hence, we can quantify changes in probability for regulatory activity attributable to given SNPs. For each regulatory function, we would expect to see a significant positive correlation between the effect-size of causal alternate alleles on cis-gene expression (beta-coefficient) and the change in probability of their association with the regulatory sequence cluster. In this way, we identified multiple regulatory sequence clusters including those with promoter or enhancer activity in myeloid and T cell lineages (Figure 2D; Supplementary Table S4).

To explore the cellular context in which the lead cis-eQTL SNPs operated we undertook co-localisation analysis within a ±200bp window centred on each lead SNP, across publicly available eQTL catalogues derived from diverse cell and tissue types. In this analysis, we sought to identify context-specific eQTL datasets which showed significant enrichment for the TST cis-eQTL. The majority of significantly enriched datasets were derived from monocytes, but also included a selection of T cell datasets (Figure 3A). Since the statistical power to identify response eQTL is partly dependent on the magnitude of response, we reasoned that myeloid and T cell lineages in the TST would show significantly greater expression levels of genes regulated by our catalogue of cis-eQTL in the TST transcriptome, compared to TST response genes for which no eQTL had been identified. We tested this hypothesis by comparing the average expression of modules of genes with and without cis-QTL associations stratified by TST single cell transcriptomes from an independent cohort^13^. We found significantly increased expression levels of TST eGene modules exclusively in two distinct myeloid lineage cell clusters differentiated in the original analysis on the basis of selected genes with antimicrobial or antigen presenting function (Figure 3B). Taken together, these analyses provide orthogonal evidence that our catalogue of cis-eQTL principally relate to transcriptional regulation in myeloid cells.

**Figure 3.**
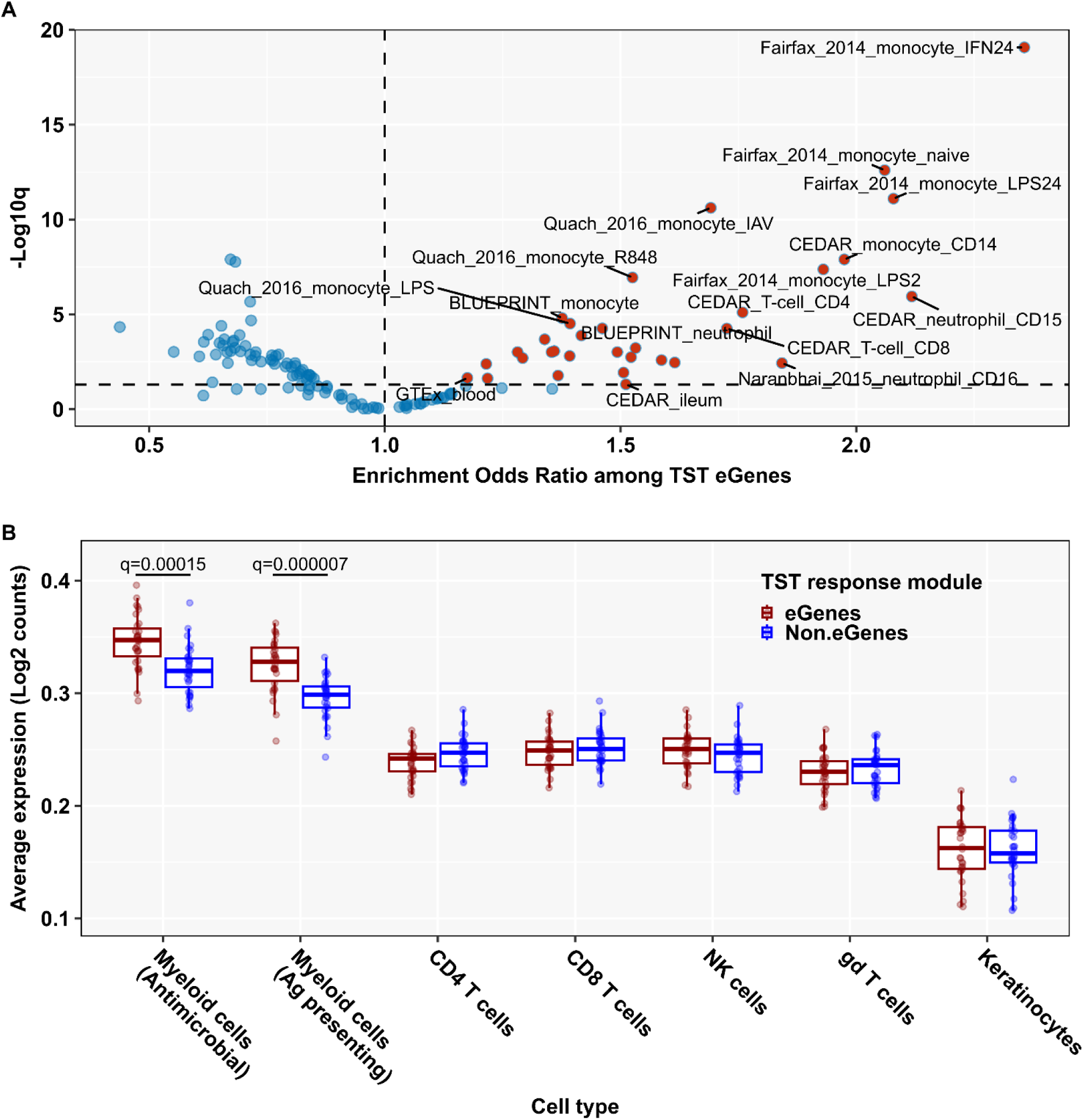
Cellular context of TST response cis-QTL. **(A)** False discovery rate (FDR) and odds ratio for enrichment of TST cis-QTL colocalization with cis-QTL lists from published studies^8^. Labels indicate abbreviated name of selected studies from which each previously published cis-QTL list is derived to include the cellular or tissue origin of the study. **(B)** Log2 transformed gene expression counts in single-cell transcriptomes from the TST^13^, averaged across genes associated with a TST response cis-QTL (shown in red), or averaged across TST response genes for which no cis-QTL was identified in the present study (shown in blue). FDR values shown for statistically significant differences (q<0.05) identified by Wilcoxon signed rank test.

### Co-localisation of TST eQTLs with GWAS disease susceptibility loci

eQTL provide the opportunity to relate disease susceptibility loci identified by GWAS to functional effects on gene expression through co-localisation of associated SNPs. GWAS analyses in TB are yet to identify consistent disease susceptibility loci in independent studies which satisfy adjusted p-value thresholds for genome-wide multiple testing^7^. However, in addition to the application of TST to model immunity to Mtb, TST responses may model generalisable cell mediated immunity. Therefore, we explored the extent to which TST cis-acting eQTL may explain heritability of disease traits in published GWAS data using partitioned heritability analysis in which the QTL were divided into two groups depending on whether their cis-gene was part of the TST transcriptional response. This showed significant enrichment in the heritability explained by TST response eQTLs vs non TST response eQTLs across a wide range of immunological diseases (Figure 4A). We then explored the specific overlap of TST response cis-acting QTLs with GWAS data in Crohn’s disease as a prototypic immunological disease which shares the feature of granulomatous inflammation with TB, and for which numerous genome-wide susceptibility loci have been established^19^. We found 17 SNPs between the two data sets with >75% posterior probability for colocalization, and statistically significant associations with susceptibility to Crohn’s disease (Figure 4B; Supplementary Table S5). Instances in which the direction of the effect of the disease allele was concordant with its effect on cis-gene expression (eg disease OR and eQTL beta coefficient were both positive or were both negative) suggests increased gene expression contributes to disease susceptibility. The strongest such association in Crohn’s disease was evident for *ERAP2*. Alternatively, disease alleles with a discordant effect on cis-gene expression suggests reduced gene expression contributes to disease susceptibility. The strongest such associations in Crohn’s disease were for *RNASET2* and *ARFRP1*.

**Figure 4.**
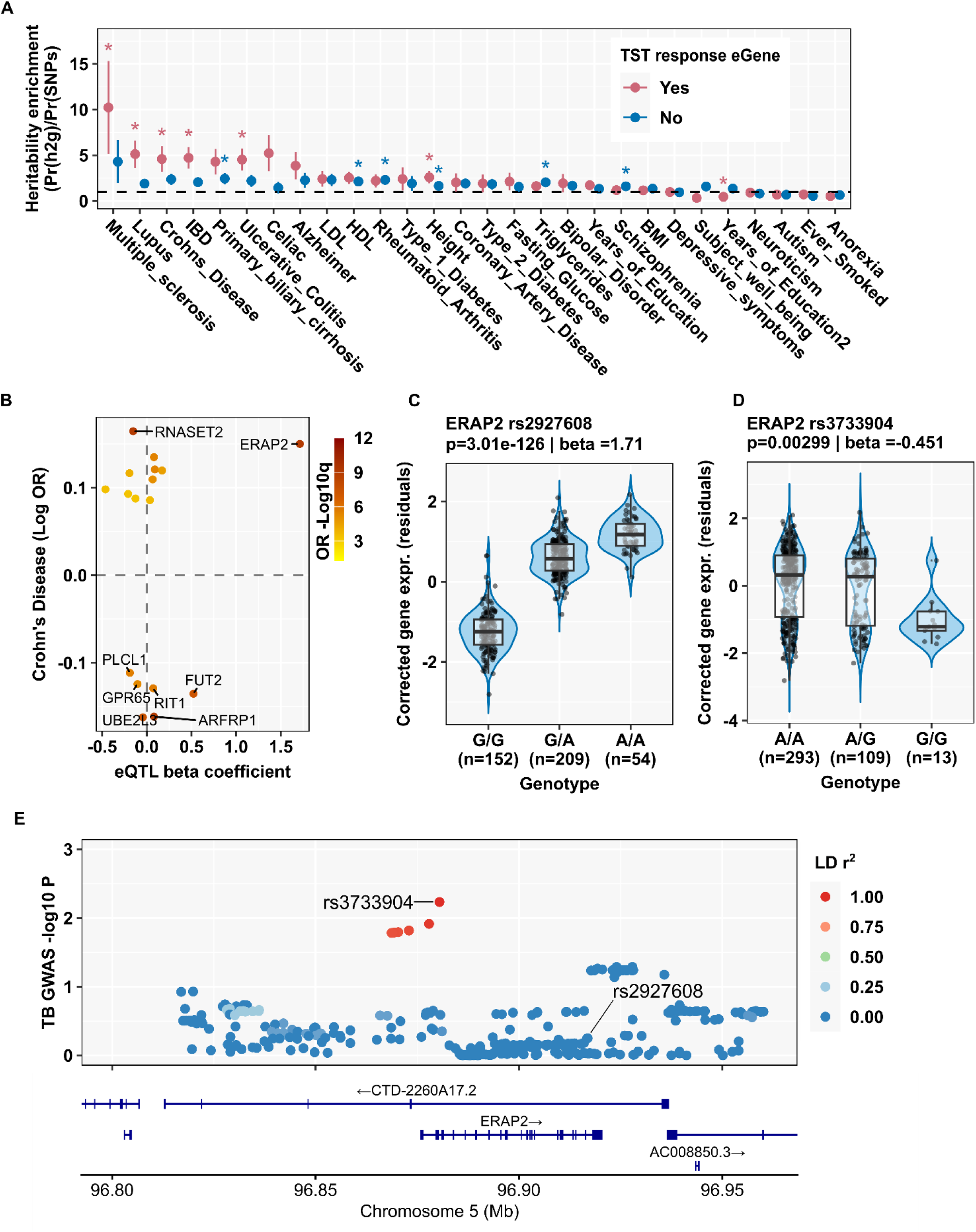
Partitioned heritability of GWAS disease risk associated with TST response genes and TST response QTL colocalisation with Crohn’s disease and TB GWAS. **(A)** Enrichment of partitioned heritability of GWAS disease traits for cis-QTL of TST response eGenes compared to cis-QTL of all other eGenes in the TST. Data show point estimates and jackknife standard errors^54^. *denotes statistically significant enrichment with false discovery rate (FDR)<0.05. **(B)** Log odds ratio of disease risk (stratified by FDR) and eQTL beta coefficients for eGenes with colocalised lead SNPs in the TST in Crohn’s disease GWAS data. **(C-D)** *ERAP2* Log2 expression residuals from multivariable analysis of the integrated TST dataset stratified by associated lead cis-QTL, rs2927608 (C) and alternative cis-QTL, rs3733904 in LD with the lead cis-QTL (D), showing the associated QTL p value and beta coefficient. (E) Regional association plot for the chromosome 5 ERAP2 locus SNPs risk of TB in multi-ancestry analysis highlighting ERAP2 cis-QTLs rs2927608 and rs3733904.

Polymorphisms in the *ERAP2* locus are of particular interest because they have been reported to have opposing effects on autoimmune/inflammatory disease and infectious disease^20^. Therefore, we undertook a targeted analysis of our lead eQTL for *ERAP2* (rs2927608, ch5:96252432:G:A) in TB. The alternate allele at this locus is associated with increased *ERAP2* gene expression and increased risk of Crohn’s disease (Figure 4B-C), but we found no signal for this SNP in differential risk of incident TB within a multi-ancestry meta-analysis of published TB GWAS data^7^ (Figure 4E, Figure 5). We did find statistically significant TB risk polymorphisms within the *ERAP2* locus, centred on rs3733904 (ch5:96216173:A:G) (Figure 4E). Interestingly, this locus is in strong LD (Lewontin’s D-prime value of 0.92) with our lead TST *ERAP2* eQTL (rs2927608) amongst Europeans specifically (Supplementary Figure 6). rs3733904 is also an eQTL for *ERAP2* gene expression within the TST, in which the alternate allele is associated with reduced *ERAP2* gene expression, but increased TB risk across multiple studies (Figure 4D, Figure 5). These findings are consistent with the general hypothesis that *ERAP2* gene expression is positively associated with autoimmune/inflammatory disease and extends the inverse association with infectious disease risk to include TB.

**Figure 5.**
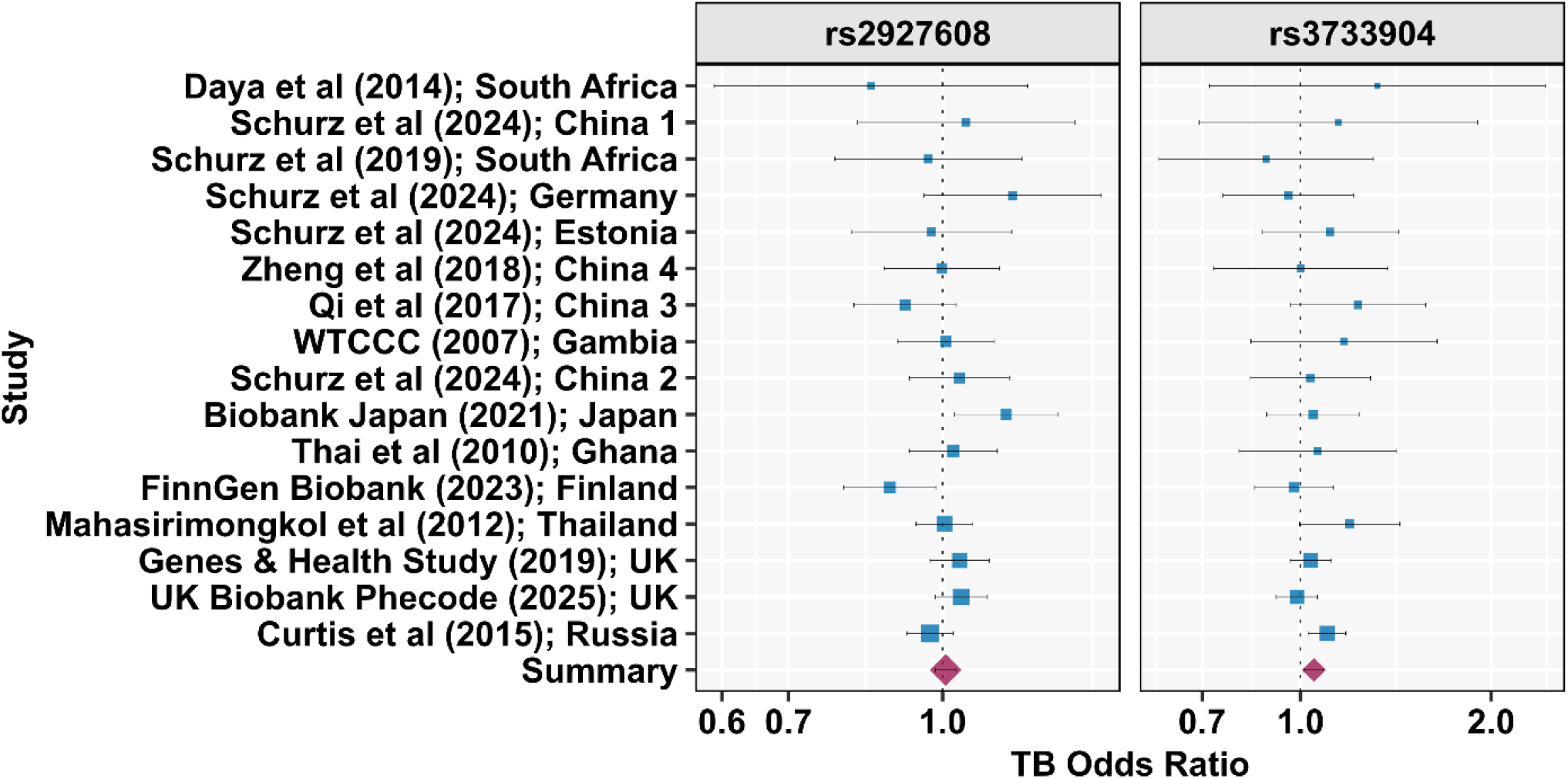
ERAP2 SNP relationship to TB disease risk Forest plot of TB odds ratios for linked ERAP2 cis-QTL in a multi-ancestry fixed-effects meta-analysis. Data show inverse variance (point size) and 95% confidence intervals, stratified by study and summary results.

### QTL regulation of antigen presentation and Mtb-reactive T cell responses in the TST

Next, we sought to use our eQTL analysis to identify the most heritable immune response pathways in the TST. We first identified Reactome pathways that were enriched among 1719 TST eGenes. The constituent genes for each significantly enriched pathway were then used as a gene-set, which we used to perform leading edge gene-set enrichment analysis in which all TST response eGenes were ranked by the statistical strength of their cis-QTL association as a proxy for heritability. Ten Reactome pathways generated gene-sets that showed statistically enriched heritability by this measure (Figure 6A, Supplementary Table S6). Seven of these ten Reactome pathways related to T cell stimulation. Since Reactome pathways and therefore our gene-sets were not comprised of mutually exclusive genes, we reasoned that the leading-edge analysis may be driven by a common set of genes shared by each of the enriched pathway gene sets. A graphical network representation of the associations between all enriched pathway gene-sets and all their constituent genes revealed multiple HLA class 2 genes as part of multiple gene sets (Figure 6B). *HLA-DRB1* was notable among this list of HLA class 2 genes because we have recently identified the most public Mtb-reactive T cell metaclones (polyclonal T cells which share a cognate peptide-MHC) in D7 TSTs to be HLA-DRB1 restricted^14^. The alternate haplotype represented by the lead eQTL was associated with a highly significant negative beta coefficient for *HLA-DRB1* expression (Figure 6C). Importantly, DRB1-restricted metaclones exhibited inter-individual variation in D7 TST frequencies due to variation in their expansion over time in the TST (Figure 6D-E). Since this T cell response is dependent on antigen presentation, we tested the hypothesis that the expression levels of *HLA-DRB1* in D2 TSTs may determine variation in subsequent antigen-specific T cell responses in D7 TSTs. Among individuals who were homozygous for their *HLA-DRB1* allele, we found a statistically significant correlation between *HLA-DRB1* gene expression in D2 TSTs and DRB1-restricted Mtb T cell reactivity in D7 TSTs (Figure 6F). To extend this analysis, we sought to identify the specific DRB1-allele restricted T cell responses that may be genetically determined by cis-eQTL for DRB1 gene expression, thereby functioning as metaclone (mc)QTL. In this analysis, we investigated the correlation between beta coefficients for the effect of cis-acting SNPs on DRB1 expression and beta coefficients for their effect on allele-specific metaclone frequencies at D7 (Figure 6G, Supplementary Figure 7A). We found a significant positive correlation for most DRB1 alleles, consistent with the hypothesis that genetically encoded levels of HLA class 2 expression may contribute to the magnitude of T cell responses. Importantly however, there were also examples in which the QTL effect on gene expression and metaclone expansion were inversely correlated. This favoured the alternative hypothesis that the associations between DRB1 expression QTLs and DRB1-allele specific T cell responses arise from linkage to coding variants that regulate peptide binding specificity. Consistent with this hypothesis, we found distinct SNP QTL for specific DRB1-allele restricted T cell metaclone expansion (Supplementary Figure 7B).

**Figure 6.**
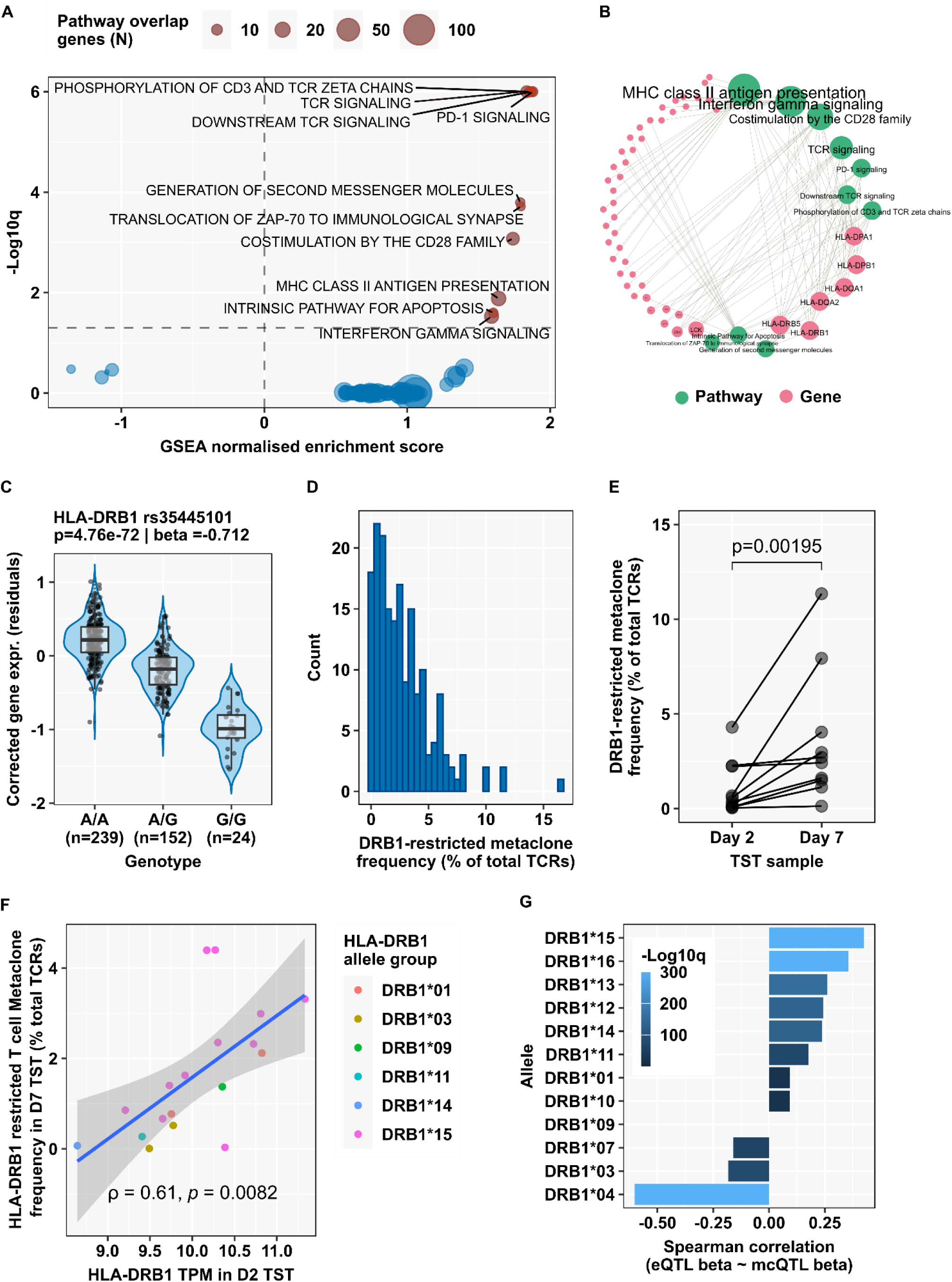
HLA class II cis-QTL and relationship with HLA-restricted Mtb-reactive T cell responses in the TST. **(A)** False discovery rate and leading edge gene set enrichment analysis (GSEA) scores for heritability of Reactome pathway gene sets that are significantly over-represented in TST response cis-QTL eGenes. Labels show selected Reactome pathways with FDR <0.05 for GSEA normalised enrichment scores. **(B)** Circular network plot of Reactome pathways (green nodes) with significantly enriched heritability of their constituent genes (pink nodes). Node/label size and order of nodes (clockwise) ranked by number of interacting edges. **(C)** HLA-DRB1 Log2 expression residuals from multivariable analysis of the integrated TST dataset stratified by associated lead cis-QTL showing the associated p value and beta coefficient. **(D)** Frequency distribution of HLA-DRB1 restricted T cell metaclones from n=178 published TST bulk TCR repertoires^14^. **(E)** Comparison of HLA-DRB1 associated T cell metaclone frequency between day 2 and day 7 TST within n=10 individual participants (p value derived from Wilcoxon signed rank test). **(F)** Relationship between day 2 TST HLA-DRB1 gene expression (Log2 transformed transcripts per million) and day 7 TST HLA-DRB1 associated T cell metaclone frequency for all HLA-DRB1 homozygous participants (n=18), showing individual data points (stratified in colour by HLA-DRB1 allelic group), linear regression line, correlation coefficient and Spearman rank test p value. **(G)** Spearman correlation between beta coefficients for HLA-DRB1 eQTL (quantifying genetic effect on HLA-DRB1 gene expression in day 2 TST) and HLA-DRB1 associated T cell metaclone (mc)QTL (quantifying genetic effect on frequency of HLA-DRB1 associated T cell metaclones in day 7 TST), stratified by HLA-DRB1 allelic group. The false discovery rate (FDR) for each association is shown by colour.

### QTL cell cycle regulation as a determinant of T cell responses

Identification of trans-acting eQTL with >5 megabase distance from the TSS of an associated gene has been used to reveal distal enhancer sequences that may exert a direct effect on gene expression through 3-dimensional chromosomal folding, or indirectly via the action of cis-associated genes. We reasoned that trans-acting eQTL may reveal either contemporaneous co-regulated biological processes within D2 or D7 TSTs, or D2 gene expression that influences downstream biology at D7. We performed trans-eQTL mapping using the same covariates as in the cis analysis, restricting tests to lead cis-eQTLs identified at D2 or D7 and to SNP–gene pairs located >5 Mb from the gene TSS. Analyses across D2 and D7 transcriptomes revealed markedly more trans associations when D7 cis-eQTLs were tested in D7 samples. Trans-eGenes were significantly enriched among TST response genes at D7 (136 of 246) compared to D2 (10 of 42) (p=0.000187; OR=3.94). Reassuringly, we found the fewest number of trans-associations between D7 cis-QTLs and D2 gene expression (Figure 7A, and Supplementary Figure 8; Supplementary Table S7).

**Figure 7.**
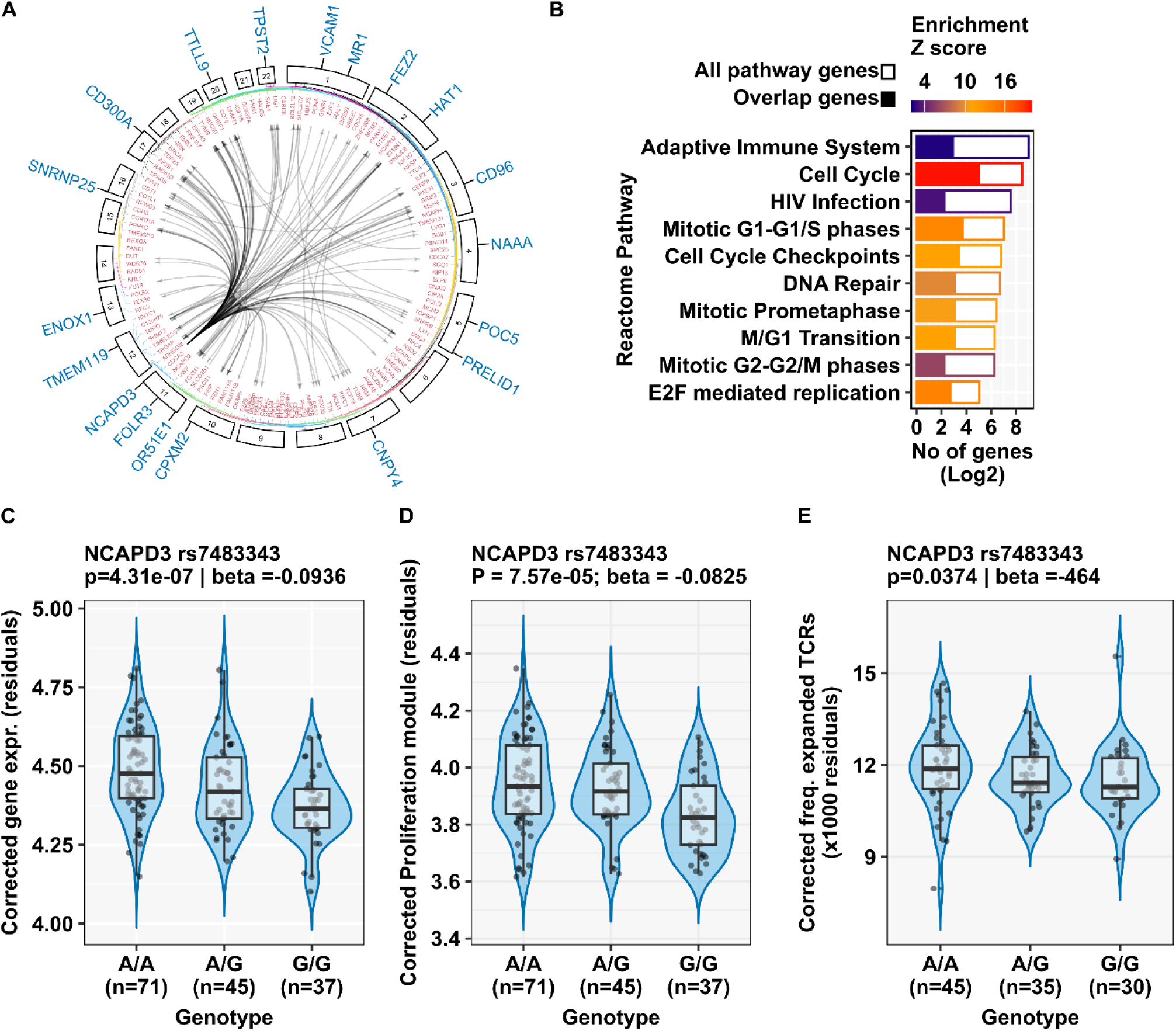
Trans-QTL associations with cell-cycle genes and T cell proliferation in the TST. **(A)** Circos plot of QTLs for cis eGenes (blue outer layer) and trans eGenes (red inner layer) in day 7 TST, stratified by chromosome location. **(B)** Reactome pathway enrichment Z scores for trans eGenes associated with the cis-eQTL for NCAPD3. **(C)** NCAPD3 Log2 expression, **(D)** cell proliferation transcriptional module and **(E)** expanded (>1 clone count) TCR frequency residuals from multivariable analysis of D7 TST stratified by the lead cis-eQTL for NCAPD3 showing the associated p values and beta coefficients.

Most D7 cis-trans associations were driven by a single cis-eQTL at NCAPD3 (81.6%; 111 of 136 trans-associations). *NCAPD3* encodes a subunit of the Condensin II protein complex involved in chromosome segregation and organisation during mitosis^21^. The trans-associated genes were also highly enriched for genes associated with cell cycle regulation and mitosis (Figure 7B), suggesting common genetic variation as a potential determinant of cellular proliferation. Accordingly, we found a significant association of the same *NCAPD3* allele with expression of an independently derived cell-proliferation transcriptional signature (Figure 7C-D). We have previously related cellular proliferation with enrichment of Mtb-reactive T cells in the D7 TST^14^. In the present analysis, we also found a statistically significant association between the lead cis-acting SNP for *NCAPD3* and frequency of expanded T cells clones (TCR count>1) in D7 TST data (Figure 7E), indicative of antigen independent mechanism for variation in antigen-specific cellular immunity. To further validate these findings in independent data, we took advantage of a recently published single cell eQTL study of T cells with and without TCR stimulation using anti-CD3/CD28^22^. We found the same cis-eQTL association for *NCAPD3* in 5day stimulated HLA-DR positive T effector memory subset (Supplementary Figure 9).

## Discussion

We present to our knowledge, the first eQTL analysis of a dynamic *in vivo* human immune response to a standardised experimental challenge that models cell mediated immunity to Mtb using the TST. We identify a large catalogue of common SNPs that show statistically significant associations with expression levels of cis-genes in multivariable analyses. We show evidence for variation in T cell responses mediated by an antigen-specific mechanism through genetic variation in the HLA-locus, and variation in T cell responses by an antigen-independent mechanism through genetic polymorphisms that regulate expression of cell cycle genes, mitosis and cellular proliferation. In GWAS data, our catalogue of TST response QTLs shows significant enrichment for the heritability of numerous immunological diseases. Finally, we show proof of concept for their application to identify functionally relevant GWAS disease susceptibility loci, and their effect on gene expression in data sets from TB and Crohn’s disease.

Our study was made possible by the finding of striking inter-individual variation in the expression levels of individual genes, suggesting regulation of transcriptional responses independent of IFNγ-producing Th1 responses that are thought to be the canonical driver of the TST^23–25^. Therefore, the TST provides a valuable model to investigate immune variation independent of IFNγ-signalling which we know is not sufficient for protective immunity to TB. SNPs are evident throughout the genome and cis-QTL are expected for all genes^26^, albeit their functional impact will in some cases be limited to specific contexts reflecting cell and cell-state specific epigenetic profiles. We identified cis-QTLs for approximately one third of TST response genes reflecting the limitations of statistical power in our sample size. Therefore, our catalogue of cis-QTL was biased towards those with higher alternate allele frequency and greater effect size. Interestingly, it was also conspicuously enriched for cis-QTL identified in independent data sets derived from monocytes. This was consistent with our observation in single cell sequencing data from the TST that expression of cis-(e)genes was significantly enriched in myeloid cell clusters, from which we infer that the dynamic nature of transcriptional regulation in this context makes the monocyte/macrophage cell lineage particularly sensitive to immune response variation by gene regulatory polymorphisms. To discriminate between causal and linked SNPs, we used a well-established probabilistic model to identify the lead SNP associated with variation in gene expression, and leveraged a large-scale foundational model of experimental functional genomics to decipher the potential gene mechanisms by which our cis-QTL may affect transcription. This analysis revealed multiple categories of regulatory function for which the effects of our QTL catalogue on cis-gene expression correlated with the probability of their modulation of promoter and enhancer sequences. Experimental verification of causal effects for individual cis-QTL is still required for specific QTLs and cis-genes of interest.

The wider application of our catalogue of cis-QTLs for cell mediated immune responses in the TST is to revisit accumulating GWAS of infectious and immunological disease-risk and provide mechanistic insights afforded by the effect of these SNPs on their cis-genes. Greater heritability of wide-ranging immunological diseases in GWAS catalogues attributable to the TST response QTLs underscores the value of our generalisable model for cell mediated immunity. Co-localisation analysis with GWAS data for Crohn’s disease showcased the opportunity to link disease-risk to functional modulation of gene expression. This was exemplified by the association between increased *ERAP2* gene expression and Crohn’s disease-risk consistent with the wider literature linking ERAP2 function with immunological and inflammatory disease, likely mediated by its canonical function as an aminopeptidase in the HLA class I antigen processing pathway leading to increased antigen presentation^17^. Although GWAS have failed to show consistent disease risk loci for TB with genome-wide statistical significance, the reported positive associations of *ERAP2* expression levels with protection against *Yersinia pestis*^20^, motivated us to evaluate its role in TB. This led to the identification of an *ERAP2* eQTL associated with reduced gene expression and increased risk of TB. Higher *ERAP2* expression may plausibly contribute to protection against TB by increasing HLA class I antigen presentation and consequently CD8 T cell immunity. Importantly, however, *ERAP2* expression also increases cell-intrinsic restriction of *Y. pestis* by macrophages, suggesting non-canonical antimicrobial effects of ERAP2 independent of antigen presentation and CD8 T cell immunity^20^. Whether *ERAP2* expression also contributes to macrophage restriction of Mtb is not known and is a priority in future work.

CD4 T cell immunity, dependent on antigen presentation by HLA class II, is necessary for protection against TB^2,3^. In our analysis, cis-QTL for HLA class II genes underpinned identification of variation in MHC class II antigen presentation and T cell activation pathways as the most heritable systems in the TST. We have previously shown that D7 TSTs are highly enriched for clonally expanded Mtb-reactive TCR sequences, from which we identified Mtb-reactive T cell metaclones representing clusters of similar TCR sequences across individuals who show enrichment for specific HLA-allelic groups. Of note, the most generalisable Mtb-reactive T cell responses were HLA-DRB1 restricted^14^. Our finding in the present study, of a correlation between variation in HLA-DRB1 expression levels in the D2 TST and subsequent expansion of HLA-DRB1 restricted metaclones in D7 TSTs, is consistent with the hypothesis that genetically encoded regulation of HLA class II expression levels may influence HLA-restricted T cell responses independently of variation in HLA restriction of antigenic binding. This was further supported by significant positive correlations between the effects of cis-QTL on expression of HLA-DRB1 and frequency of DRB1-allele specific metaclone QTLs. However, these data do not resolve the possibility of linkage between non-coding QTLs and coding sequence variation that restricts antigen binding. Indeed, examples of significant negative correlations between QTL effects on gene expression and T cell metaclones can only be explained by linkage in which restriction of immunodominant antigens mediates the relationship between HLA and T cell responses. Of note, the TST cis-QTL for HLA-DR did not colocalise with the statistically strongest HLA-DR associated loci in the TB GWAS meta-analysis^7^.

Our analysis of trans-QTL associations provided novel biological insight into an alternative antigen-independent mechanism for genetic regulation of T cell responses, mediated by cell proliferation through QTL effects on genes that control cell cycle and mitosis. Future work will need to investigate whether a cis-acting QTL for *NCAPD3* exerts wide-ranging trans-effects on cell cycle genes by acting as a direct enhancer, via the effects of chromatin condensation on transcriptional regulation^27^, or via an entirely novel function of NCAPD3 independent of its role in chromatin condensation^28^. Importantly, we identified the same cis-QTL effect on *NCAPD3* in a subset of activated T cells in an independent data set. These findings highlight a previously neglected mechanism for heritable variation in immune responses. NCAPD3 has been identified as a potential oncogenic factor in a range of solid organ and haematological cancers^29–35^. Increased NCAPD3 expression has also been associated with tumour immune cell infiltration^36^, immunopathology of ulcerative colitis^37^ and protective immunity to bacterial infection^38^. Experimental validation of SNP regulation of *NCPAD3* expression would provide invaluable Mendelian randomisation instruments to further evaluate the causal associations of NCAPD3 with infectious and immunological diseases.

Sample size limitations precluded analyses stratified by disease state for robust identification of differential eQTL associations in LTBI and ATB. However, our combined analysis increased statistical power and provided generaliseable results across the spectrum of Mtb infection and TB disease. Further extension of the sample size in this model is required to obtain comprehensive coverage of genetic regulation of immune response variation to leverage the maximum value of GWAS data. The mixed ancestry of our study generates additional challenges to statistical power, but also affords the advantage of greater generalisability of results to date. The application of bulk transcriptomics required indirect approaches to infer the cellular context in which the identified QTLs operate. These inferences will require future experimental validation alongside SNP editing experiments to identify the causal SNPs that regulate gene expression of interest. Nonetheless, our findings have extended validation of diverse QTL previously reported in laboratory models to complex multi-cellular immune responses. We have identified a novel mechanism for regulation of adaptive T cell responses by QTL that regulate cell proliferation and a novel association between genetically encoded levels of *ERAP2* gene expression and TB risk for investigation in future mechanistic studies.

## Methods

### Study approvals

Research ethics and regulatory approvals for the present study were provided by UK research ethics committees (reference numbers: 16/LO/0776, and 18/LO/0680). All study participants provided written informed consent.

### Study population and sampling

Study participants comprised two separate cohorts as previously described^10^ (Table 1, Supplementary Figure 1). The first cohort sampled patients with presumed latent TB infection (LTBI). These were healthy HIV seronegative adults, with immune memory for Mtb-specific antigens identified by positive peripheral blood IFNγ release assays using the QuantiFERON Gold Plus Test, but no clinical or radiological evidence of active tuberculosis disease. The second cohort sampled patients with active TB disease (ATB) within 4 weeks of starting TB antimicrobial treatment. All participants received TSTs comprising 2U intradermal tuberculin (Serum Statens Institute) in the volar aspect of the forearm as previously described^9,10,12^. The precise injection sites were marked with permanent marker pen and used to position the site of the TST punch biopsies at designated time points. LTBI participants underwent a TST in both arms. One site was biopsied on D2, and the second site was biopsied on D7. The ATB cohort underwent a single TST that was biopsied on D2. In each case, 3mm punch biopsies from the injection sites were cryopreserved in RNALater (Qiagen) and used to extract total RNA as previously described^10^. All participants also provided a peripheral blood sample in EDTA used to extract genomic DNA using the QIAamp spin column (Qiagen). TST biopsy data from these two patient cohorts was compared to that of previously reported biopsies from the site of saline injections^10^ to identify TST response genes (described below).

### RNA sequencing and analysis

Total RNA from TSTs were subjected to genome wide mRNA sequencing as previously described^10^.

For QTL analysis, sequencing reads were trimmed using Trim Galore (v0.6.2) and aligned to the human genome (hg19) using HISAT2 (v2.1.0)^39^, followed by gene-level read counting with featureCounts (v1.6.2) using GENCODE v31 annotations. Only genes with transcript-per-million (TPM) values ≥ 1 in at least 10% of samples were retained, giving 25,886 genes in D2 LTBI samples, 26,364 genes in D7 LTBI samples, and 25,900 genes in ATB samples.

For all other analyses, sequencing reads were pseudo-aligned using Kallisto^40^ with the Ensembl Human GRCh38 release 111 transcriptome. Transcript-level counts were summed on gene level, and annotated with Ensembl gene ID, gene symbol and gene biotype using the R/Bioconductor packages tximport and BioMart. Ensembl gene IDs, retained after exclusion of pseudogenes, were used for differential expression analysis with the SARtools (v1.8.1) implementation of DeSeq2^41^, with a false discovery rate (FDR) <0.05 and log2 fold difference of ≥0.58 (fold change ≥1.5). TPM values were log2 transformed after the addition of a pseudocount of 0.001. Duplicated gene symbols were filtered by retaining the gene with highest expression per sample. Principal component analysis was performed using the prcomp function in R. Spearman correlation coefficients were used to assess the relationship between IFNγ gene expression and (i) TST principal components, or (ii) individual TST gene expression variance (calculated as standard deviation across individuals). Venn diagram analysis of the TST transcriptomes in each data set was conducted in DeepVenn^42^.

Upstream regulator analysis of differentially expressed genes was performed using Ingenuity Pathway Analysis (Qiagen). This was visualised as a network diagram using the Force Atlas 2 algorithm^43^ in Gephi v0.9.4 and used to derive co-regulated gene-expression networks as previously described^44^. The analysis was restricted to upstream regulators predicted to be significantly activated (Z-score>2, adjusted p-value<0.05), targeting at least 4 downstream genes, and annotated with one of the following functions: cytokine, kinase, transmembrane receptor, and transcriptional regulator, representing the canonical components of pathways which execute transcriptional reprogramming in immune responses. For each upstream regulator, pairwise Spearman correlations of the TPM expression values of the target genes were calculated among TST samples. Upstream regulators were selected as significant if the average co-correlation was significantly (FDR <0.05) greater than the distribution of average correlation coefficients obtained from 100 iterations of selecting an equivalent number of random genes. Reactome pathway enrichment of TST response eGenes or trans QTL-associated genes was analysed with the XGR (v1.1.9) R package^45^.

Gene set enrichment analysis (GSEA) was performed with GSEA v4.3.3^46^ using the GSEA Preranked tool with the 1719 TST response eGenes ranked by eQTL q-value (as a surrogate for heritability) as the input gene list. The 84 Reactome pathways which were enriched in the overall gene-list were used as gene-sets, in order to identify which pathways were enriched in the most heritable genes and to identify which highly heritable genes comprised the enrichment leading-edge. Recommended GSEA default parameters were used (1000 gene-set permutations and FDR q-value <0.05). Pathways with GSEA FDR <0.05 were visualised as network plot using the circular layout algorithm^43^ in Gephi v0.9.4.

The transcriptional module for cellular proliferation has been derived and published previously^44^, and module gene expression was quantified as the arithmetic mean log2 TPM value of its constituent genes.

### T cell receptor (TCR) repertoire analysis of TSTs

Processed bulk TCR sequencing data from day 2 and day 7 TST biopsies of the LTBI cohort were published previously^14^ and are available from UCL’s Research Data Repository [https://doi.org/10.5522/04/28049606]. Frequency of expanded beta-chain TCR clones was calculated as abundance of TCRs with count >1.

HLA-DRB1-associated Mtb-reactive T cell metaclones are groups of similar beta-chain TCR clones shared across day 7 TSTs from different individuals with significantly enriched HLA-DRB1 allele expression^14^. Metaclone abundance was quantified by comparing the V gene and CDR3 amino acid sequence of beta-chain TCRs to the V gene and CDR3 regular expression of each pre-defined metaclone. The frequency of HLA-DRB1-associated TCR metaclones in TST repertoires was calculated as percentage of all TST TCRs matching a metaclone description. To correlate HLA-DRB1 gene expression in day 2 TSTs with metaclone frequency in day 7 TSTs, the abundance of TCR metaclones associated with a specified HLA-DRB1 allele was quantified in individuals homozygous for that HLA-DRB1 allele. To assess the genetic effect on the frequency of HLA-DRB1 associated T cell metaclones in day 7 TSTs, the abundance of TCR metaclones associated with a specified HLA-DRB1 allele was quantified in all individuals expressing that allele, regardless of their zygosity.

### Genotype data processing and imputation

Genome-wide genotypes at 1,904,599 loci were generated across 267 individuals using the Illumina Infinium Global Diversity Array-8 v1.0 (Supplementary Figure 3A). Raw intensity idat files were converted to genotype call gtc files using Illumina’s iaap-cli tool, and subsequently converted to VCF format using the bcftools +gtc2vcf plugin. SNP quality control (QC) was performed using BCFtools (v1.10.2). We excluded variants located on sex chromosomes and mitochondrial DNA, retained only biallelic SNPs, and applied the following filters: minor allele frequency (MAF) ≥ 5%, SNP call rate ≥ 95%, and SNP-only sites, giving a total of 677,069 autosomal SNPs after QC. Pre-phasing was performed using Eagle (v2.4), and genotype imputation was carried out with Minimac4 (v1.0.2) via the Michigan Imputation Server 2. The Haplotype Reference Consortium (HRC) r1.1 panel was used as the reference, selected for its slightly improved imputation performance relative to the 1000 Genomes Phase 3 panel (Supplementary Figure 3B). After imputation, SNPs were retained if they met the following criteria: imputation quality score (R²) ≥ 0.8, MAF ≥ 5%, and Hardy-Weinberg equilibrium P value ≥ 1e-05 in at least one of the five defined populations. The number of SNPs passing all filters and used in the downstream eQTL analyses were: 5,961,895 for D2 LTBI samples, 5,994,645 for D7 LTBI samples and 5,992,435 for ATB samples.

### Genotype principal component analysis

We computed genotype principal components (PCs) from imputed SNP data using PLINK^47^. To assess ancestry, we downloaded reference genotype data from the 1000 Genomes Project and identified SNPs shared between the reference panel and our TB cohort. Genotypes were pruned for linkage disequilibrium using a sliding window approach (--indep 50 5 1.5) in PLINK. Principal component analysis (PCA) revealed clustering consistent with five major ancestral populations represented in the cohort (Supplementary Figure 3C). Genotyping QC identified no related samples and all 267 individuals were retained for analysis.

### Cis-eQTL analysis

Cis-eQTL mapping for each dataset (D2 LTBI, D7 LTBI and D2 ATB) was performed using an additive linear model as implemented in R package MatrixeQTL^48^. Autosomal SNPs were tested for association with gene expression if located within ±1 Mb of the gene’s transcription start site (TSS). To control for population effects on the discovery of eQTLs, genotype principal components (PCs) were used as covariates. The first five genotype PCs captured the major population structure in this cohort, showing clear the associations between PCs and different ethnicities (Supplementary Figure 3D). To maximise power and control for hidden confounders, we included gender, age, and the first five genotype PCs, along with the optimal number of expression PCs that maximised eQTL discovery^49^. Gene expression levels for eQTL mapping were quantified as log₂-transformed TPM using an offset of 1 based on gene counts obtained as described above.

To identify significant cis-eGenes, we performed 1000 permutations using the --permute option as implemented in fastQTL5 (v2.0), with significance modelled using beta distributions. A second level of multiple-testing correction was applied across all tested genes using the Benjamini-Hochberg procedure on beta-adjusted *P* values. and threshold ≤ 0.05 to define genes with at least one significant cis-eQTL (eGenes).

We performed cis-eQTL mapping across all 415 RNA-seq samples from 267 individuals using a linear mixed model. As above, SNP-gene pairs were tested for association if the variant was located within ±1 Mb of the gene’s TSS. The model included covariates for gender, age, the first five genotype PCs, and the optimal number of gene expression PCs (Supplementary Figure 4). To account for repeated measures and relatedness, linear mixed models were fitted using the R package *lmerTest*^16^. We computed the P values based on Satterthwaite’s degrees of freedom method as implemented in *lmerTest*. The model was specified as follows: 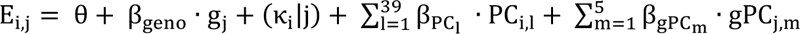, where E_i,j_ is gene expression for the ith sample from the jth individual, *θ* is an intercept term, and *β*_geno_ is the eQTL effect of the genotype for individual j (g_j_). (κ_i_|j) is the random effect accounting for variability between individuals. We included the optimal 39 gene expression PCs (β_PC_l_) and 5 genotyping PCs (β_gPC_m_) as fixed effects as described above.

To define the significant cis-eGenes, we used the eigenMT pipeline^50^ to estimate the effective number of independent tests (Meff) per gene and applied Bonferroni correction to compute locally adjusted p-values for each gene’s peak association. A second level of multiple-testing correction for the adjusted p-values was then applied across all genes using the q-value function in R, and threshold ≤ 0.05 to define eGenes as above.

### Conditionally independent cis-QTL mapping

To identify conditionally independent cis-eQTLs, we used an iterative two-stage mapping approach consisting of forward regression followed by backward selection as previously described^51^. To do this, we first calculated a gene-level p-value threshold corresponding to a q-value of 0.05 by identifying p-values near the boundary of significance. Specifically, we identified the smallest adjusted p-value (Bonferroni-corrected using the effective number of tests, *Meff*, based on eigenMT as described above) among genes with q-values >0.05, and the largest adjusted p-value among those with q-values <0.05. The global threshold was defined as the average of these two values. A gene-level p-value threshold was then obtained by dividing the global threshold by the *Meff* for each gene.

Using this gene-level threshold, we next performed iterative cis-eQTL mapping, correcting for all previously identified lead SNPs that passed the gene-level significance threshold and the standard covariates used in the initial analysis. This forward selection process continued until no additional independent lead variants were detected. In the backward stage, each SNP was scanned for cis-QTLs individually while conditioning on all other independent lead variants found in the forward stage, as well as the standard covariates. If no significant association was found using this full model, it was dropped. Otherwise, the most significantly associated variant was selected as the lead SNP for each independent signal.

### Trans-eQTL analysis

For *trans*-eQTL mapping, we used the same covariates as for *cis*-eQTL analysis. We restricted our analysis to lead cis-eQTLs (q-value ≤ 0.05) identified in either D2 or D7 LTBI samples. We considered only SNP-gene pairs where the SNP was located more than 5 Mb from the transcription start site (TSS) of the gene. Pseudogenes were excluded for candidate trans-eGenes based on GENCODE v31 annotations. Gene-level FDRs were calculated using the Benjamini-Hochberg method, correcting p-values observed across all tested trans-genes for each cis-eQTL. We further removed false positive trans-eQTL caused by reads cross-mapping with cis regions^52^.

### Colocalisation analysis

Colocalisation analysis was performed using the R package coloc (v5.2.3)^53^ with default settings, within a ±200 bp window centred on each lead eQTL. The coloc method applies a Bayesian framework to estimate the posterior probability that signals from different eQTL datasets, or between eQTL and GWAS datasets, share a common causal variant (PP4). Colocalisation was defined as PP4 > 0.9. eQTL summary statistics were obtained from the eQTL catalogue^8^, comprising 127 datasets across 75 tissue/ell types upon 14 different treatment conditions.

### Stratified LD score regression

We used stratified LD Score Regression (S-LDSC)^54^ to estimate partitioned heritability from GWAS summary statistics based on both pre-defined functional annotation sets and the annotations derived from eQTLs identified in TB patients. Enrichment was defined as the proportion of SNP heritability explained by an annotation divided by the proportion of SNPs in this annotation. To do this, we obtained the pre-computed baseline LD scores (v2.2), the 1000 Genomes Phase 3 European SNP reference panels and GWAS summary statistics from Zenodo (https://zenodo.org/records/7768714). We generated functional annotation files and computed LD scores for each eQTL category using the make_annot.py and ldsc.py scripts provided by the LDSC software (https://github.com/bulik/ldsc). We included all cis-eQTLs with nominal p-values passed the gene-level significance threshold (corresponding to a q-value of 0.05) defined using eigenMT across 7411 eGenes, as described above. Partitioned heritability enrichment was then computed using the --h2 flag in ldsc.py with default parameters incorporating LD score regression weights based on HapMap3 SNPs (excluding the HLA region) as recommended.

### GWAS coloc and overlay

Publicly available GWAS summary statistics were obtained for tuberculosis from the International Tuberculosis Host Genetics Consortium (ITHCG)^7^ and Crohn’s disease from the International IBD Genetics Consortium^19^. Following the approached used to investigate IL-6 signalling and risk of tuberculosis^55^, the ITHCG data were supplemented with summary statistics from five additional studies: BioBank Japan (case definition: ICD A15-A16, respiratory tuberculosis)^56^; Pan-UK Biobank (case definition: self-reported tuberculosis)^57^; FinnGen (case definition: ICD A15, bacteriologically and histologically confirmed tuberculosis); Genes & Health (case definition: Genes & Health custom phenotype based on ICD-10 and SNOMED codes)^58,59^; Zheng et al. (case definition: microbiologically confirmed pulmonary tuberculosis)^60^. For each dataset, we extracted chromosome, position, variant ID, alleles and standard error. Multi-allelic variants and sex chromosomes were excluded. Allele alignment between GWAS and eQTL variants was verified, with mismatched variants flagged and corrected. Fixed-effects meta-analysis was performed using the R package rmeta and metasoft^61^. Files were sorted, compressed with bgzip, and indexed using tabix for efficient querying. For each trait-associated locus (±200 kb around the lead eQTL variant), we extracted summary statistics for shared GWAS and eQTL SNPs, and performed colocalisation analysis using coloc with default settings as described above. Estimates of linkage disequilibrium were obtained from the 1000 Genomes Project using LDlink^62,63^.

### Data and code availability

For the LTBI cohort, RNAseq data have been deposited in the European Genome-phenome Archive (EGA) under accession number EGAD50000001208. For the ATB cohort, RNAseq data have been deposited in Array Express (accession number: E-MTAB-6816). Genotyping data have been deposited in EGA (accession number EGAD00010002811) and include a metadata table that allows linking of genotype and RNAseq IDs. All code used for data processing and analysis is publicly available on GitHub (https://github.com/jknightlab/HIRV-TB_eQTL, and https://github.com/carolinturner/tst_eqtl).

## Data Availability

https://ega-archive.org/datasets/EGAD50000001208

https://www.ebi.ac.uk/biostudies/arrayexpress/studies/E-MTAB-6816

https://ega-archive.org/datasets/EGAD50000000288

## Acknowledgements

This work was supported by Wellcome Trust awards 207511/Z/17/Z and 306550/Z/23/Z to MN, 204969/Z/16/Z to JCK and 222098/Z/20/Z to TP. MN and MB also acknowledges support from NIHR Biomedical Research Funding to University College London Hospitals. JCK acknowledges support from NIHR Oxford Biomedical Research Centre and from the Medical Research Council (MR/V002503/1). PZ and JCK acknowledge support from the Chinese Academy of Medical Sciences (CAMS) Innovation Fund for Medical Science (CIFMS), China (2024-I2M-2-001-1). Computation used the Oxford Biomedical Research Computing (BMRC) facility, a joint development between the Wellcome Centre for Human Genetics and the Big Data Institute, supported by Health Data Research UK and the NIHR Oxford Biomedical Research Centre. GST acknowledges support from a Medical Research Council Clinician Scientist Fellowship (MR/N007727/1). We thank Victoria Dean, Michelle Berin, Zandile Maseko and Kimberlee Gunn, for supporting participant recruitment and sampling. For the purpose of open access, the authors have applied a CC BY public copyright licence to any Author Accepted Manuscript version arising from this submission.

## Author contributions

Conceived and designed the study: GST, JCK, MN.

Sample and clinical data collection: AC, GST, JR, RRK, MB, SC, ML, HK, SL.

Laboratory analysis: AC, JR.

Data analysis: PZ, CT, TP, LCKB, JJ, JCK, MN

Manuscript preparation: PZ, CT, TP, JCK, MN with input from all authors

