## Supplementary Figures for "Common genetic determinants of dynamic human in vivo immune response to *Mycobacterium tuberculosis*"

**Supplementary Figure 1. Consort diagram of study groups and procedures**

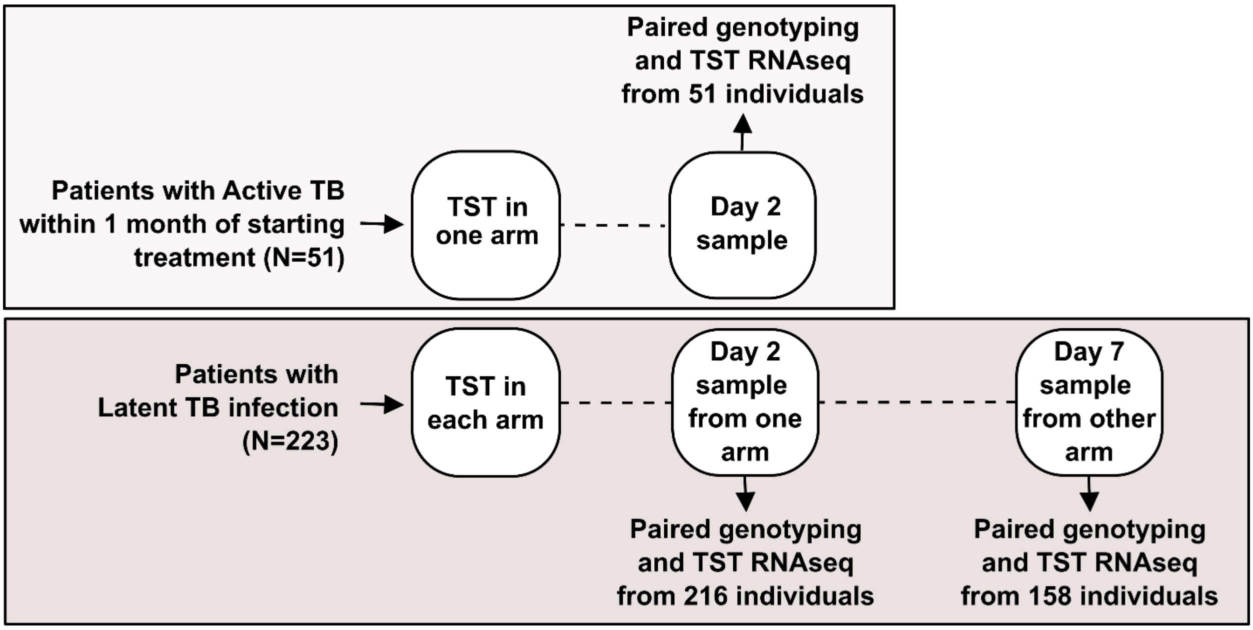

**Supplementary Figure 2. TST transcriptome correlation with *IFNG* expression**

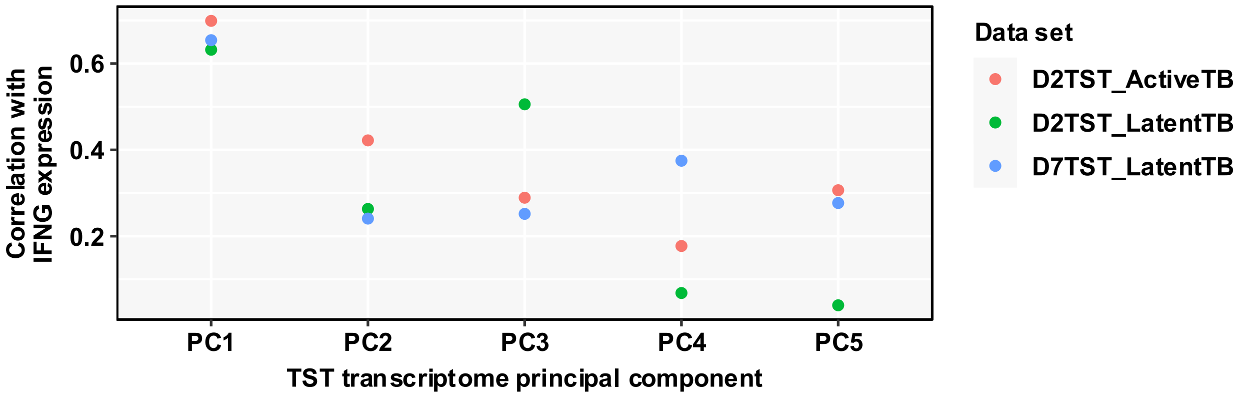

Spearman correlation coefficients of TST transcriptome principal components (PC) and *IFNG* gene expression in the TST, stratified by dataset.

**Supplementary Figure 3. Genotype QC and imputation.**

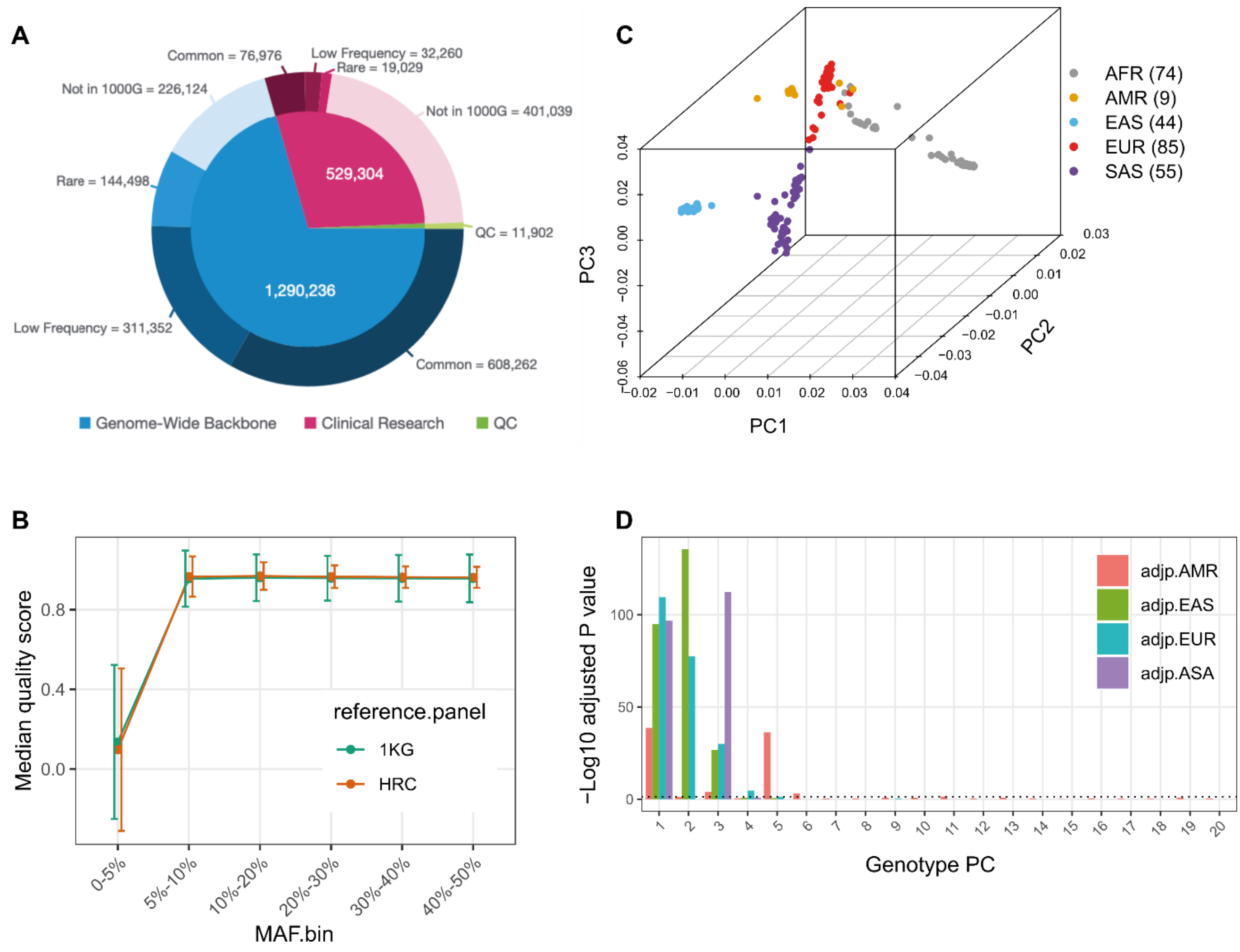

**(A)** Genotyping loci on the Illumina Infinium Global Diversity Array-8 v1.0. **(B)** Principal component analysis (PCA) of genotypes derived from 267 TB patients. **(C)** SNP imputation results based on the 1000 Genomes Project (green) and Haplotype Reference Consortium (HRC) r1.1 (red) panels, showing the median imputation quality score ($R^2$) on the y-axis across different minor allele frequency (MAF) bins. **(D)** Bar plot showing the association between a given genotype PC (x axis) and different ethnicities (African as a reference) using linear regression. the FDR-adjusted P value was shown on the y axis (−log10 scale).

**Supplementary Figure 4. Cumulative cis-eQTL associations by principal components included in additive linear models**

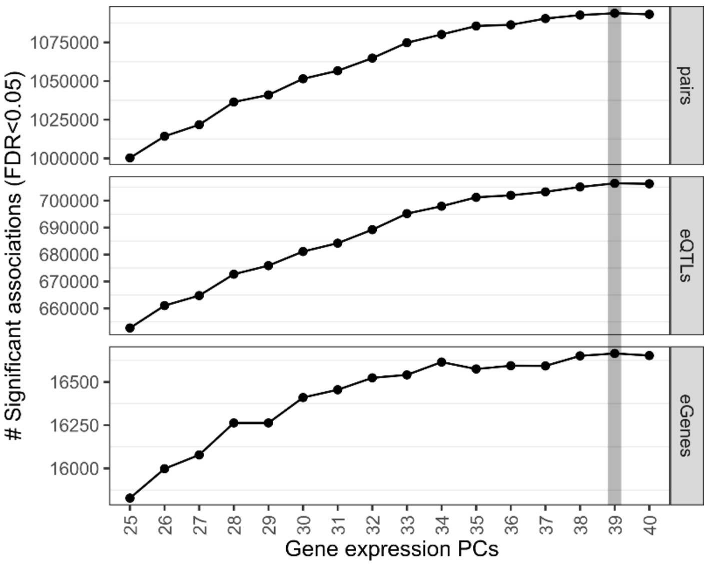

Line plot showing the number of significant cis-eQTL associations (FDR< 0.05, y-axis) across different numbers of gene expression PCs included in the linear mixed model.

**Supplementary Figure 5. Cis-eQTL count stratified by effect size and alternate allele frequency**

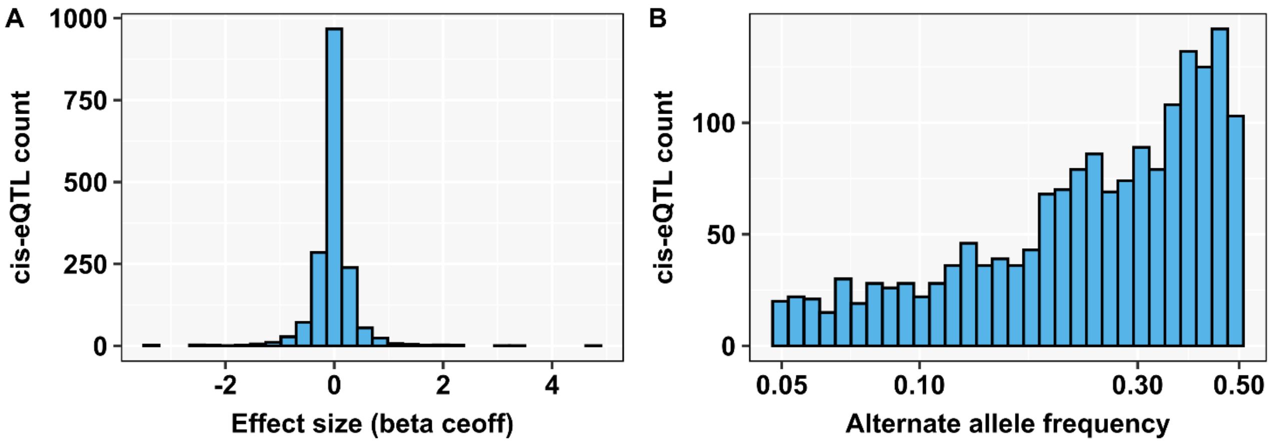

Frequency of TST response cis-QTL stratified by cis-eQTL beta co-efficient as a measure of effect size **(A)** or alternate allele frequency **(B)**.

**Supplementary Figure 6. Linkage disequilibrium of SNPs in ERAP2 locus across global and European ancestries.**

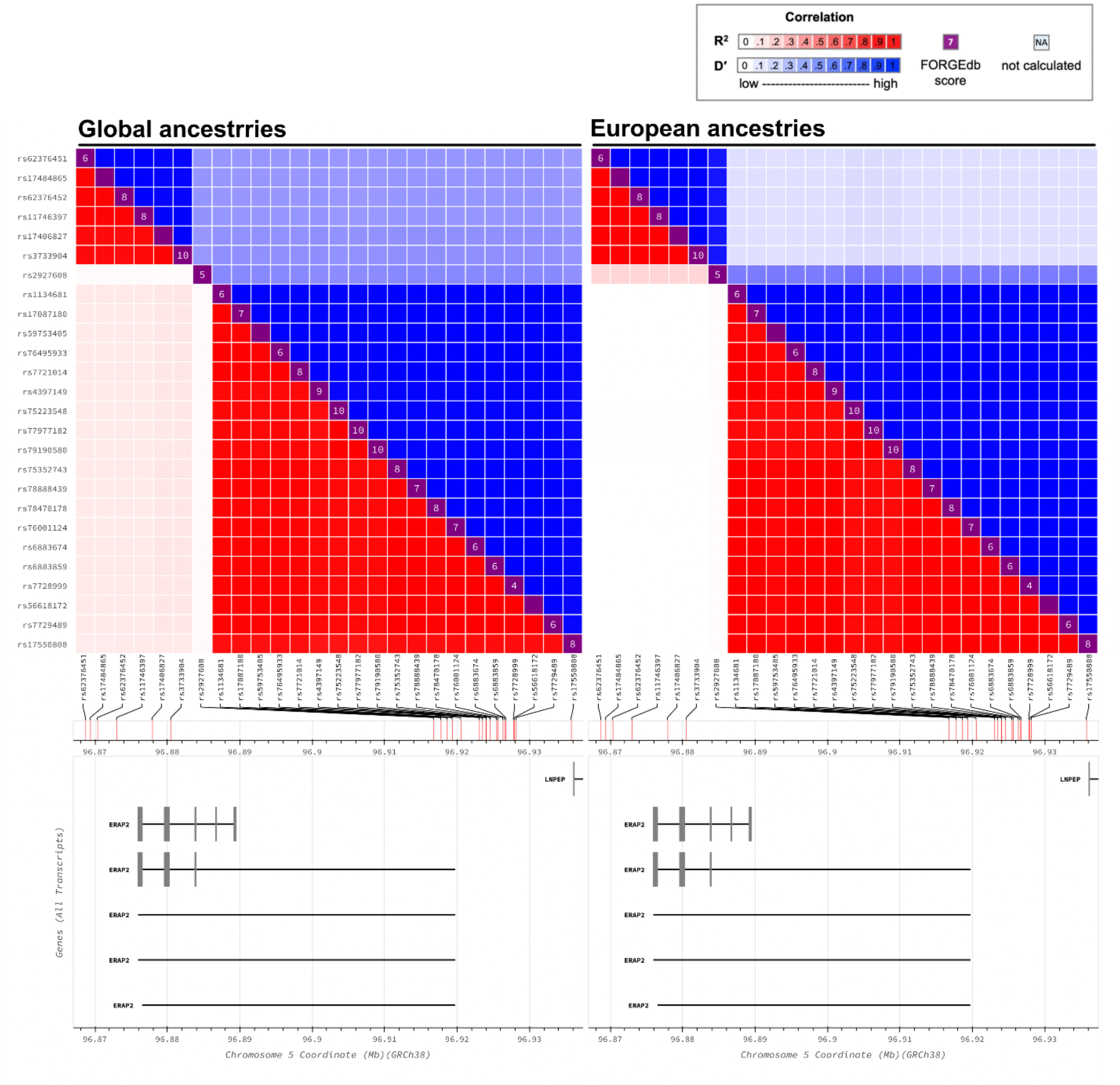

Heatmap matrix of pairwise linkage disequilibrium statistics prepared using the LDmatrix tool (ldlink.nih.gov)[1]. Variants other than rs2927608 were included if the p-value from the multi-ancestry fixed-effects meta-analysis for susceptibility to TB disease was less than 0.1. Linkage disequilibrium is indicated using a colour gradient from white to red ($R^2$=1) and blue (D'=1). Numbers in purple squares on the diagonal are FORGEdb scores which indicate the likelihood that a variant is regulatory ranging from 0 to 10 (forgedb.cancer.gov)[2].

**Supplementary Figure 7. HLA allele QTL of gene expression and HLA-restricted Mtb-reactive T cell responses in the TST.**

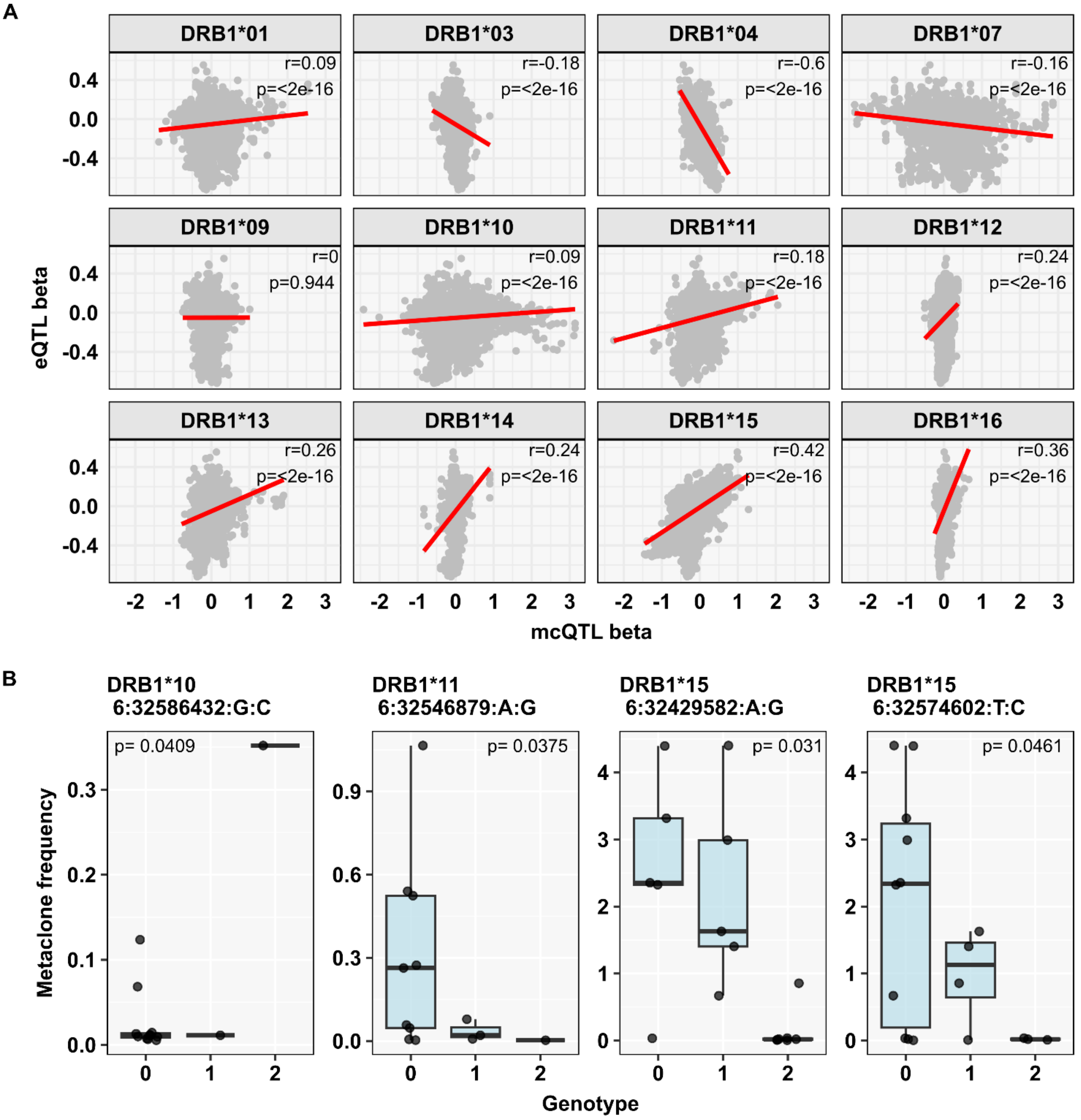

**(A)** Relationship between HLA QTL effect size on gene expression (e) in integrated TST data and effect size on HLA-restricted Mtb-reactive T cell metaclone (mc) frequency in day 7 TST data, with linear regression lines in red and Pearson's correlation coefficient/p-value indicated. **(B)** Lead mcQTL for HLA-allele restricted day 7 TST metaclone frequency.

**Supplementary Figure 8. Trans-eQTL association in day 2 or day 7 TST among day 2 or day 7 TST cis-eQTL**

**A D2-cis ~ D2-trans**

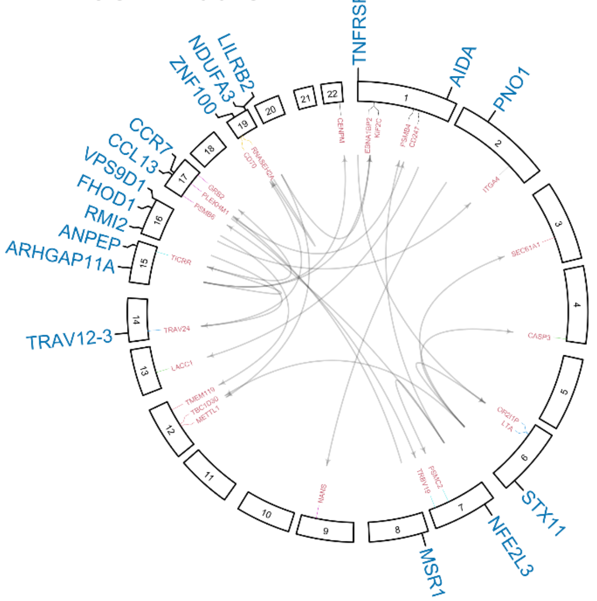

**B D2-cis ~ D7-trans**

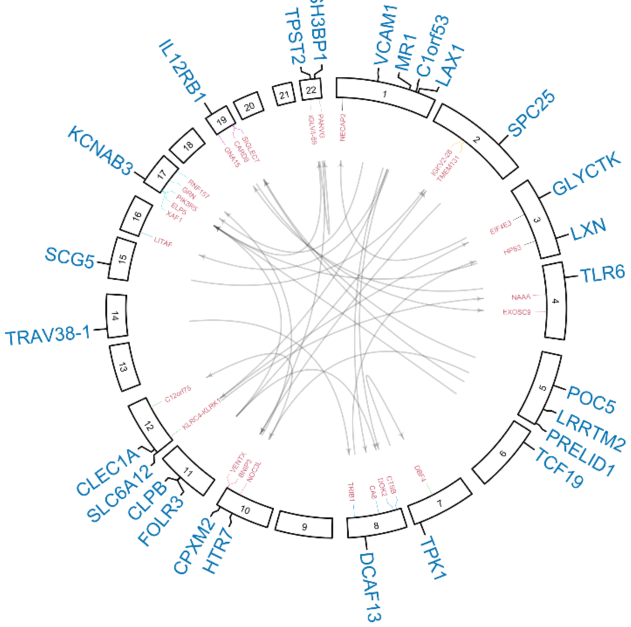

**C D7-cis ~ D2-trans**

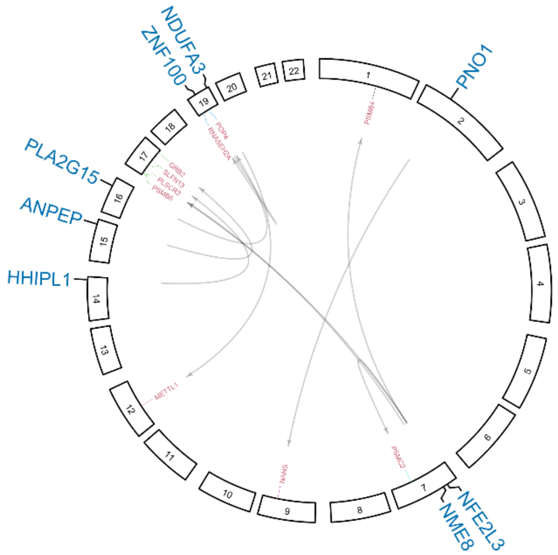

Circos plots of QTLs for cis eGenes (blue outer layer) and trans eGenes (red inner layer) for **(A-B)** day 2 or **(C)** day 7 cis-eQTL, stratified by chromosome location.

**Supplementary Figure 9. Effect of rs748334 on NCAPD3 expression stratified by cell type and time point ±TCR/CD28 co-stimulation.**

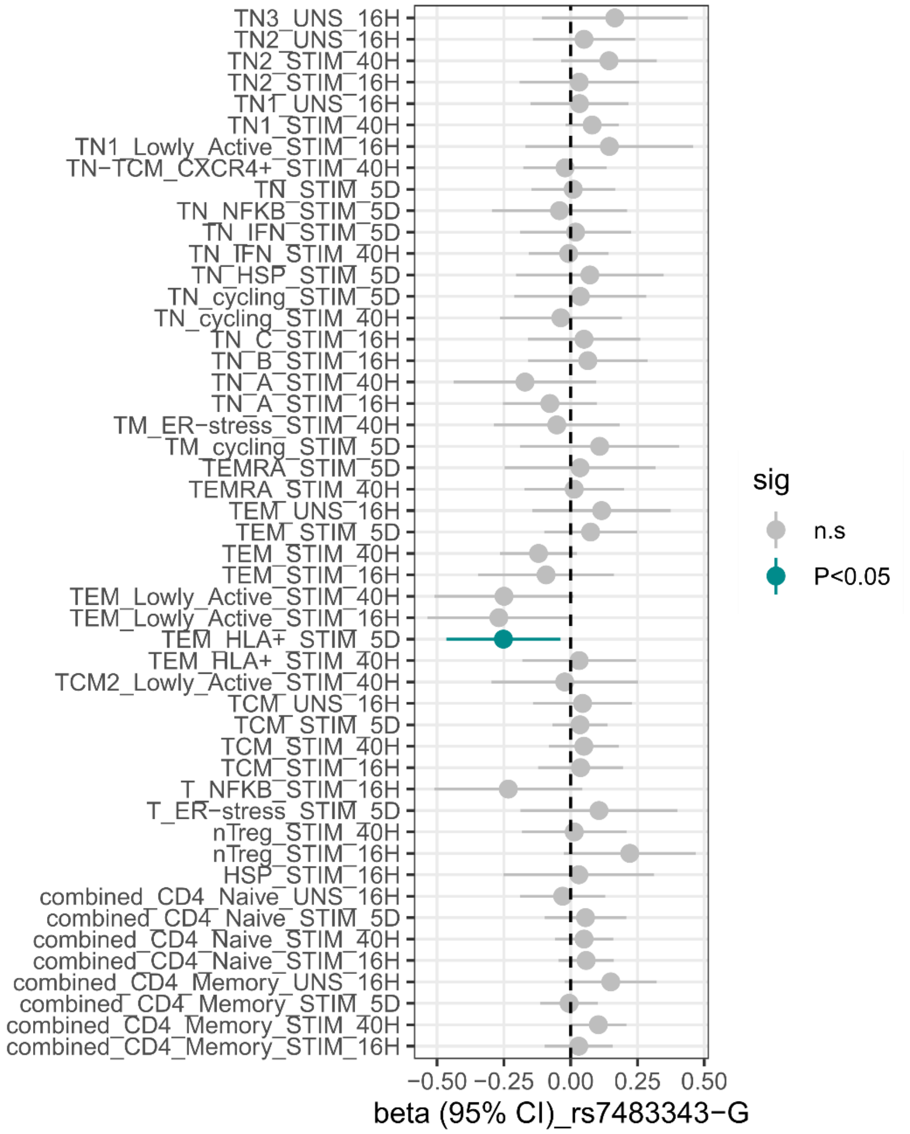

Summary of rs748334 effect (beta coefficient and 95% confidence interval) on NCAPD3 gene expression stratified by cell type ± anti-CD3/CD28 stimulation at 16-120 hours.
